# Reporting of RACE in Australian brain and mental health research: protocol for a scoping review and diversity audit

**DOI:** 10.1101/2025.08.08.25333336

**Authors:** John Noel Viana, Ben Albert Steward, Darren Suryawijaya Ong

## Abstract

**Introduction:** The underrepresentation of racial and ethnic minorities in health research in multicultural societies can lead to limited understanding of health challenges and conditions that they face. This leads to interventions that may have reduced efficacy or undocumented adverse effects on certain populations. Focusing on Australian research on post-traumatic stress disorder (PTSD), depression, stroke, and dementia in 2020-2024, this scoping review and diversity audit aims to: 1.) evaluate reporting of variables related to RACE (race; ancestry; country of birth; country of residence and migration status; culture, including language and religion; and ethnicity) and 2.) examine representation of participants from various backgrounds.

**Methods and analysis:** This scoping review will follow the Preferred Reporting Items for Systematic Reviews and Meta-Analyses – Scoping Review Extension (PRISMA-ScR) guidelines. Six scientific and medical databases (Medline, Scopus, Embase, Web of Science, PsycInfo, and CINAHL) were searched for relevant articles published from 1 January 2020 to 31 December 2024. The search retrieved 21,927 articles, and after removing duplicates via EndNote, 10,033 articles were uploaded to Covidence for screening. All titles, abstracts, and full texts will be screened in independent duplicate. Relevant data will be extracted using a template focused on diversity and demographic-related information. Findings will be reported through narrative synthesis.

**Ethics and dissemination:** Ethics approval is not required for this review. Research findings will be disseminated through a peer-reviewed publication in a medical or health journal, academic and practitioner networks, and conference presentations.

**Strengths and limitations:**

- This study will be the first to simultaneously and systematically examine racial/ethnic diversity in Australian research on four brain and mental health conditions. Relevant articles will be identified using a comprehensive search strategy that aims to capture a wide range of studies.
- The scoping review will employ a rigorous approach, adhering to PRISMA-ScR guidelines and screening all studies in independent duplicate for inter-reviewer reliability.
- The synthesis is expected to identify how race, ethnicity, and related variables are reported in Australian brain and mental health research and how different minority groups are represented. Insights will inform the next phase of a research program that aims to develop pragmatic guidelines for increasing minority representation in Australian brain and mental health research.
- A limitation is that only articles in the past five years will be reviewed.

## BACKGROUND

Multiple racial and ethnic groups are often underrepresented in brain and mental health (BMH) research globally and in multicultural societies. For example, clinical trials for Alzheimer’s disease drugs in the USA have rarely included Hispanic and Black participants,^1^ and global psychiatric genomics studies primarily involve people with European ancestry.^2,3^ Tools for measuring brain activity, such as electroencephalography and functional near-infrared spectroscopy, were not designed for people with curly hair and dark skin.^4^ Psychology publications on race are rare, with most studies being led by White researchers and employing few participants of colour.^5^ Exclusion or underrepresentation of racial/ethnic minorities in BMH research has a significant cost, as treatments, healthcare policies, and research agendas may not fully address their needs.^6^ In multicultural and settler-colony countries like Australia, 29.3% of its population are overseas-born and 3.2% of Australians identify as Aboriginal and/or Torres Strait Islander,^7^ amounting to a total of 8.5 out of 26 million Australians.

These studies underscore the importance of understanding contemporary representation of various RACE-ial (race; ancestry; country of birth; country of residence and migration status; culture, including language and religion; and ethnicity) minorities in BMH research in multicultural societies. Acknowledging that understandings of race and ethnicity continue to evolve,^8^ and can significantly vary across contexts,^9^ the term RACE-ial minorities is used throughout this protocol to account for the multiple conceptions of race, ethnicity, ancestry, and other culturally and genealogically related terms. For instance, while the term “ culturally and linguistically diverse” groups is used in Australia to include migrants from non-Anglo backgrounds and Indigenous peoples, it is used inconsistently in quantitative research and have been used to refer to country of birth, language spoken at home, Indigenous status, migrant/refugee status, and/or self-defined ethnic/cultural background.^10^

As a starting point for auditing RACE-ial diversity in health research, this review focuses on Australia and examines four BMH conditions (post-traumatic stress disorder or PTSD, depression, stroke, and dementia) that significantly impact its population. Reviewing research on these conditions would enable preliminary comparisons on RACE-ial diversity across psychiatric, neurologic, and neurovascular diseases. This review is essential, given that these conditions affect at least 4 million Australians. The lifetime prevalence of PTSD in Australia is estimated to be 11%.^11^ Around 1.5 million Australians experience depression each year,^11^ with disability-adjusted life years (DALYs) estimated to be around 181,000.^12^ At least 387,000 Australians have experienced a stroke episode, with the disease causing 4.4% of all deaths in 2022^13^ and estimated DALYs of 125,000.^12^ Finally, an estimated 411,000 individuals in Australia live with dementia,^14^ resulting in 262,000 DALYs.^12^ Economically, depression is projected to cost the nation AU$4.3 billion by 2030,^15^ and dementia alone is projected to cost the country AU$25.8 billion by 2036.^16^

Despite affecting large populations globally and in Australia, few studies have examined RACE-ial diversity in BMH research. To our knowledge, no RACE-ial diversity audits have been conducted for PTSD research, despite racial disparities in PTSD prevalence and treatment, with lifetime prevalence of PTSD highest among Black populations and with minority groups being 39-61% less likely to seek treatment compared to White people.^17^ A global audit of internet-based cognitive behavioural therapy (iCBT) trials for depression found that only 27% of studies reported race and 19% reported ethnicity, with White participants overrepresented (74.9%), and minority groups underrepresented (Black, 7.6%; Asian, 5.8%; Indigenous, 0.1-0.4%).^18^ An audit of dementia prevention trials in the USA found that the majority of participants were Non-Latino White (75.4%) with minoritised groups generally underrepresented,^19^ particularly Black (4.9%), Asian (0.3%), American Indian/Alaska Native (0%), and Native Hawaiian and Pacific Islander (0%) populations. Finally, for stroke, only 60% of published trials in the USA reported the race or ethnicity of participants; however, minority representation remained low (25% Black, 9% Hispanic, 2% Asian or Pacific Islander, and <1% Native American patients).^20^ Few trials also addressed inequities in rehabilitation, recovery, and social reintegration.^21^

Among research on the four conditions in Australia, to the best of our knowledge, only dementia studies have been audited for RACE-ial reporting. A review by Low et al. in 2019 revealed that most studies do not report ethnicity data, with 44.7% of dementia clinical trials explicitly excluding non-English speakers.^22^ Cohort studies also rarely collect data on country of birth, years lived in Australia, or language spoken at home.^22^ While there have been no recent systematic audits of RACE-ial diversity in PTSD, depression, and stroke research in Australia, several studies and reports have raised concerns on disparities in minority representation in research. A national review of Australian mental health policies, research, and services in 2007 found limited research directly addressing depression in ethnic minority communities.^23^ The review found that depression research within ethnic minorities in Australia has primarily focused on recently arrived refugee populations and postnatal depression in specific linguistic groups, such as Arabic- and Vietnamese-speaking women, while longer-term immigrant communities remain understudied. A recent evaluation of the Australian National Stroke Audit programme did not identify the collection of racial or ethnicity information as a key focus area.^24^ For PTSD, there were no studies that compared prevalence across different minority ethnic groups.

### Study Rationale

Considering the lack of up-to-date research that examines RACE-ial reporting and diversity in Australian BMH studies, this scoping review will provide an update to Low’s (2019) paper on diversity in dementia research and to our knowledge, will be the first to systematically examine RACE-ial diversity in PTSD, depression, and stroke studies. The prevalence of these conditions and their impact on the quality of life of millions of Australians underscore the pressing need to examine which populations are underrepresented and thus may not fully benefit from research on these diseases. With Australia’s growing migrant population and persistent health disparities experienced by its Aboriginal and/or Torres Strait Islander populations^25-27^ and by refugees and humanitarian entrants,^28,29^ it is imperative for BMH research to increasingly, equitably, and respectfully include diverse populations. While the Black Lives Matter and anti-Asian hate protests during the COVID-19 pandemic have amplified calls for increased diversity in research,^30-35^ it remains unclear whether these movements have had an impact on RACE-ial minority representation in Australian medical research. The suspension or cancellation of DEI-related (diversity, equity, and inclusion) programs by the USA government in 2025^36,37^ and its repercussions on research across the world, including in Australia,^38^ signifies the need to audit RACE-ial diversity from a period preceding the change (articles published in 2020-2024), which can then be used to evaluate the impacts of these policies in the coming years. Finally, with the ongoing adverse effects of interpersonal and structural racism on the mental health of minoritised Australians,^39,40^ it is crucial to examine the extent to which biomedical and health research contributes to promoting good mental health and well-being for marginalised groups.

Findings from this scoping review will inform a larger study that aims to develop pragmatic recommendations for equitably improving RACE-ial diversity in Australian BMH research (DE240100386). This scoping review is the first phase of the project, and authors from studies included in the review will be invited to participate in surveys, interviews, and workshops to investigate their perspectives on and actions towards improving RACE-ial diversity in their research. Findings from the different project phases, including the scoping review, will inform the development of an anti-racist neuroethics framework to address epistemic injustices in Australian BMH research. Examining underrepresentation through an epistemic injustice lens^6,41-43^ can facilitate a more comprehensive understanding of its impact on biomedical knowledge, knowledge production systems, and health equity. Anti-racist neuroethics also underscores the need for situated and reflexive understandings of race^44^ and for localised anti-racism efforts^45^ for diversity promotion, taking into account Australia’s colonial history, demographics, scientific research and funding infrastructure, and research ethics guidelines and protocols.

### Review Objectives

Focusing on Australian research on post-traumatic stress disorder (PTSD), depression, stroke, and dementia in the past five years, this scoping review primarily aims to:

1. Evaluate reporting of RACE, including the terms, variables, and categories used
2. Examine and audit the representation of participants from various RACE-ial backgrounds In addition, the review also aims to:
3. Compare diversity across the four conditions, different types of research (genomic, psychologic, pharmaceutical, epidemiologic, etc.), different settings (among Australian states/territories), and periods/years
4. Examine whether RACE-ial minorities were engaged in the design and conduct of the study, in line with initiatives on consumer leadership^46^ and patient-public involvement^47^
5. Determine whether studies reflect on how the demographic diversity of their participants impacts research translatability Finally, information on the lead and corresponding authors will be obtained to:
6. Invite them to participate in the subsequent phases of the research project (surveys, interviews, and workshops)
7. Determine whether the RACE-ial background of the author has an influence on the RACE-ial diversity of research participants

## METHODS

This article outlines the protocol for a scoping review, which will be conducted and reported according to the PRISMA-ScR (Preferred Reporting Items for Systematic Reviews and Meta-Analyses-Scoping Review Extension) checklist. ^48^ In addition, they will be supplemented by the Joanna Briggs Institute scoping review guidelines^49^ to facilitate rigour and replicability.

### Search strategy development and retrieval of relevant articles

Previous scoping and systematic reviews on PTSD,^50,51^ depression,^52,53^ stroke,^54,55^ and dementia,^56,57^ were used to develop the base or primary search string. We also added search strings from previous scoping and systematic reviews that exclusively examined studies in Australia.^58,59^ Furthermore, with the advice of a medical research librarian at the Australian National University, we added “ AND NOT” search strings to exclude meta-analysis, reviews,^60^ and animal studies. Search strings were added to exclude animal studies.^61^ An Australian researcher with expertise in ethnic diversity in dementia research and care was consulted to refine the search string.

Ultimately, the search strategy included three key concepts,

1. Research on any of the four conditions: PTSD, depression, stroke, and dementia
2. Studies conducted in Australia and published between 2020-2024
3. Original empirical research on human populations, samples, or datasets

These concepts were used to develop the search string, with the OVID Medline search string presented in Table 1. Search strings for other databases are presented in Appendix 1.

**Table 1.**
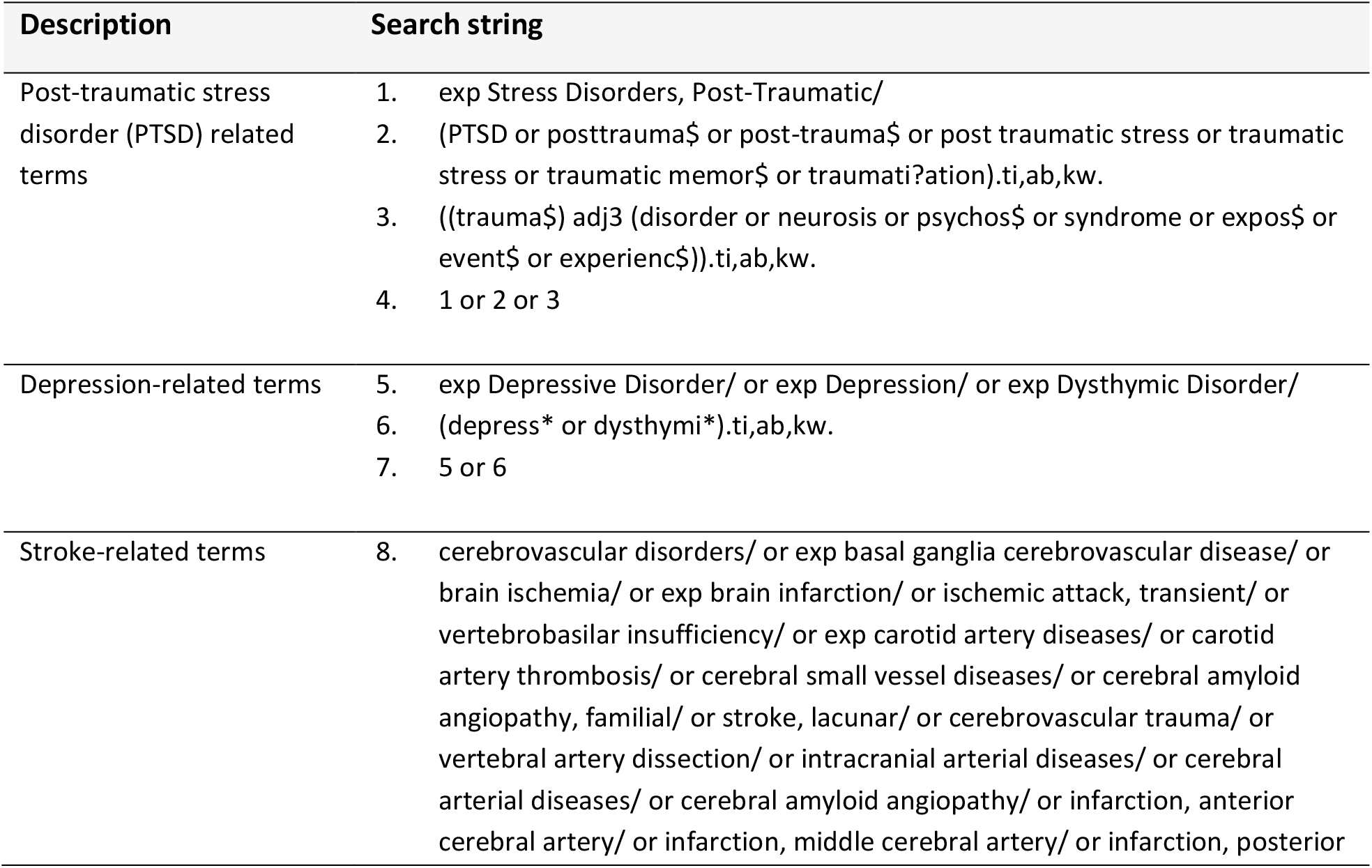
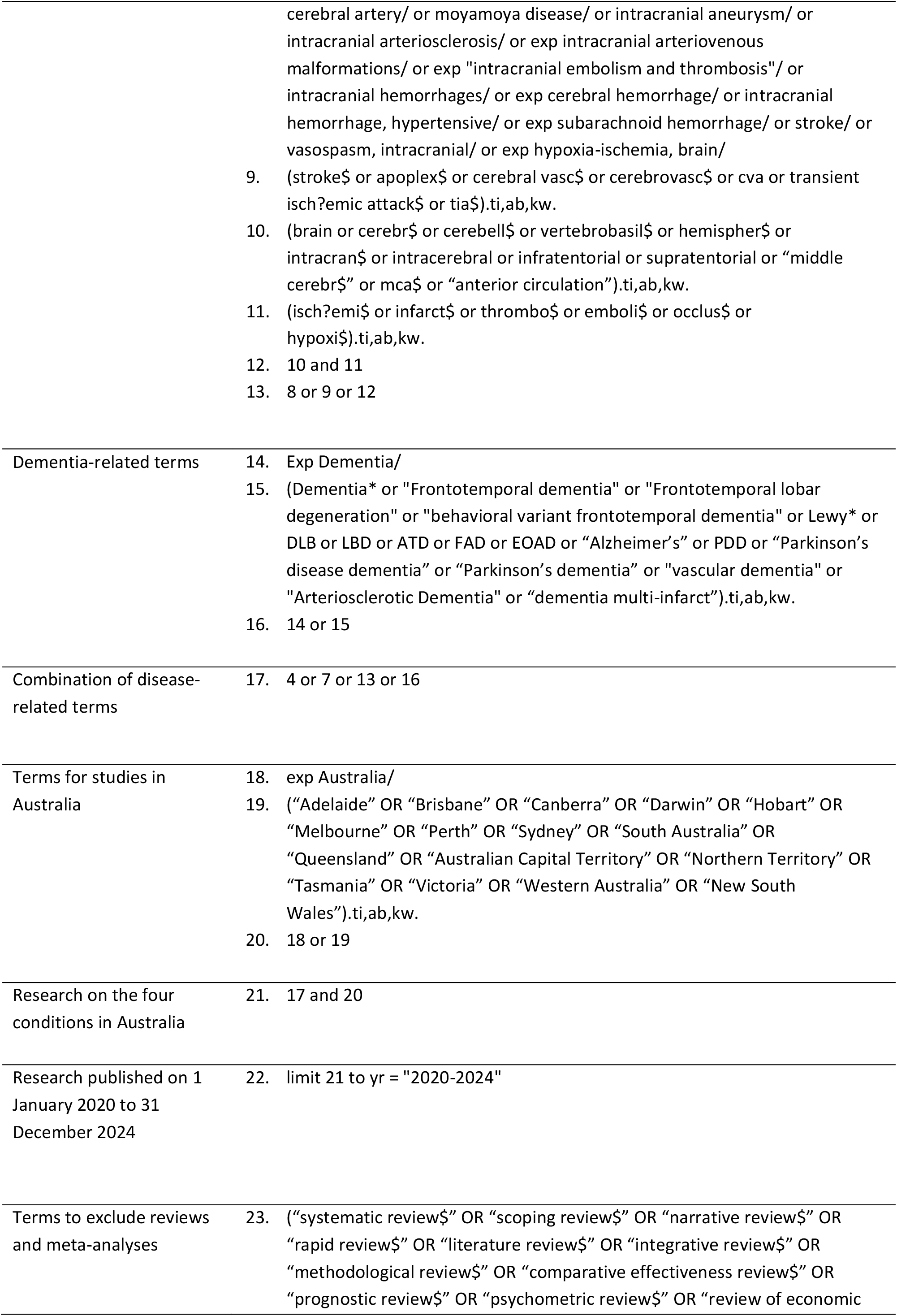
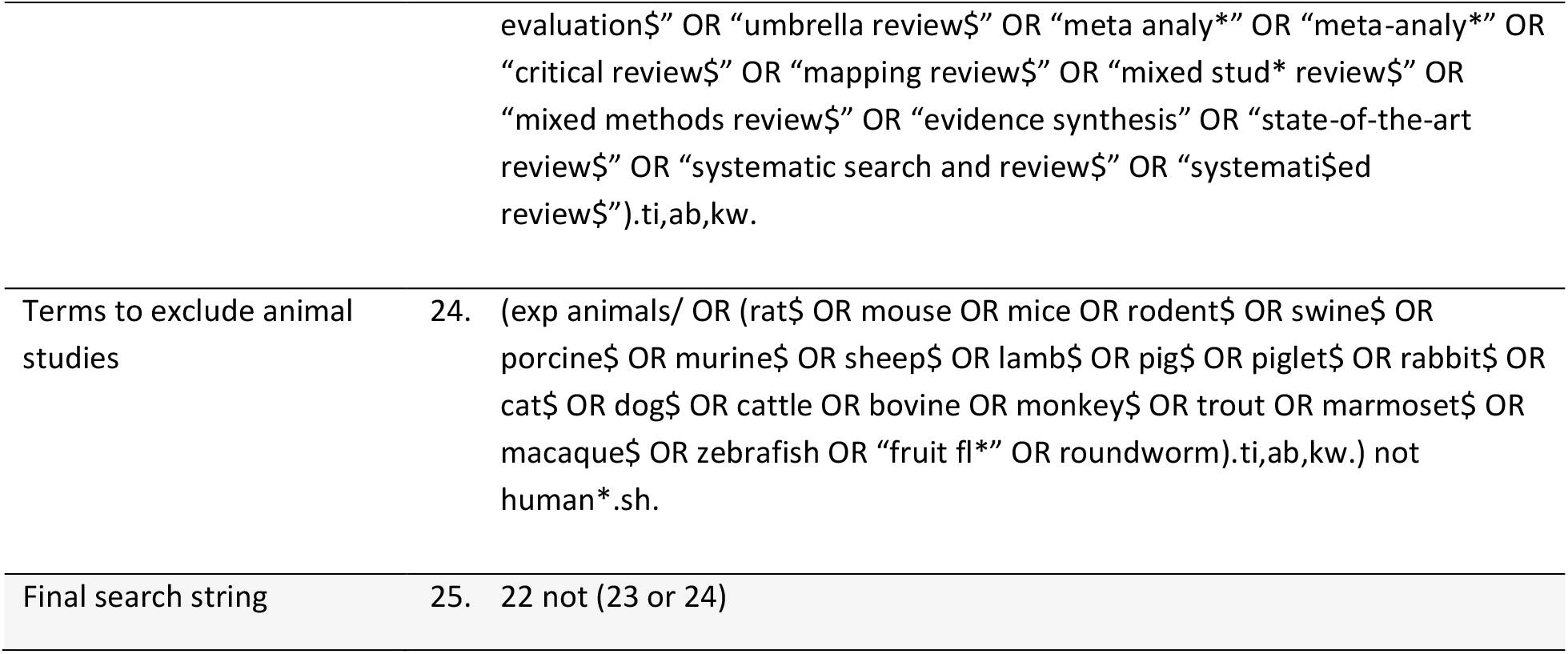
Search string for OVID Medline.

#### Information sources

Consistent with evidence regarding optimal coverage for medical and health publications,^62^ this review will retrieve articles from Medline (through OVID), Embase, Scopus, Web of Science, CINAHL, and PsycInfo.

#### Search results

We applied our search strategy across the six academic databases to retrieve peer-reviewed scientific articles. All searches were conducted on 27 March 2025. The search identified a total of 21,927 articles (Table 2).

**Table 2.** Number of articles retrieved from each database.

| Databases | Count |
| --- | --- |
| OVID Medline | 3313 |
| Web of Science | 4519 |
| Scopus | 6116 |
| Embase | 5244 |
| CINAHL | 2341 |
| PsycInfo | 394 |
| Total | 21,927 |

### Eligibility criteria and screening

Since the primary aim of the review is to determine RACE reporting in research on PTSD, depression, stroke, and dementia, few eligibility restrictions on the type of empirical study will be placed. Studies that were published between 1 January 2020 and 31 December 2024; describe original research related to the four conditions; conducted in Australia; and involve human participants, samples, and/or datasets will be included (see Table 3). Several eligibility criteria were based on previous relevant scoping reviews,^63,64^ while others were revised or added to account for the objectives of this particular review. Mock screening of 200 titles, abstracts, and full texts were conducted from 31 March to 18 April 2025 to refine and finalise the inclusion and exclusion criteria.

**Table 3.** Inclusion and exclusion criteria for title, abstract, and full text screening.

|  | Inclusion | Exclusion |
| --- | --- | --- |
| Population | Intentionally recruited human participants at risk for, with predisposition to, are diagnosed with, are experiencing, or have recovered from any of the four conditions<br>Intentionally recruited caregivers and family members of people with predisposition to, are diagnosed with, are experiencing, or have recovered from any of the four conditions<br>Lay people or non-medical/health/scientific “experts” who do not have any of the four conditions but have been evaluated for their knowledge of any of the four conditions, primary outcomes related to these conditions, or experiences with navigating the healthcare system in relation to any of the four conditions | Animal studies<br>*In silico* studies<br>Studies with no demographically identifiable human participants (commercial cell lines, geographically and demographically deidentified/detached databases, AI datasets)<br>Professional experiences or perspectives of researchers, physicians, and healthcare professionals who conduct research or provide care for people with any of the four conditions |
| Concept | For quantitative studies, targeted collection of outcomes, predictors, or measured variables related to any of the four conditions<br>For qualitative studies, one of the main interview/survey questions is related to any of the four conditions<br>*Studies where the questions or measures are just secondary or unrelated will be included if there is intentional recruitment of populations with any of the four conditions | Measures, questions, or items related to the four conditions are just treated as non-targeted outcomes or follow-up questions and there is no intentional recruitment of people with any of the four conditions |
| Condition | Any of the following:<br>1. Post-traumatic stress disorder<br>2. Depression<br>3. Stroke<br>4. Demetia | Research not related to any of the four conditions and did not include populations with any of the four conditions |
| Context | Studies conducted exclusively in Australia | Studies that are not conducted in Australia or do not indicate number of participants/samples from Australia |
|  | Multi-country studies that indicate number of participants/samples from Australia |  |
| Type of Evidence | Primary quantitative and/or qualitative empirical research (randomised controlled trials, observational studies, cohort studies, cross-sectional studies, case reports, interviews, surveys, etc.) | Protocols |
|  |  | Reviews (scoping reviews, systematic reviews, narrative reviews, umbrella reviews, rapid reviews, etc.) |
|  | Brief communications | Meta-analysis |
|  | Full text articles | Meta-ethnography |
|  | Full-text conference proceedings | Media analysis |
|  | Dissertations | Editorials, perspective pieces, or opinion statements |
|  | Articles written in English | Delphi studies, focus groups, and consensus statements from experts |
|  |  | News articles, features, and press releases |
|  |  | Economic evaluations that did not involve input from human participants |
|  |  | Conference abstracts |
|  |  | Articles not written in English |
|  |  | Articles that have unavailable or inaccessible full text |

Screening will be conducted using Covidence, a web platform dedicated for systematic reviews, and it will be performed across multiple stages. Before uploading the articles to Covidence, EndNote was used to automatically and manually remove duplicates. There were 11,894 duplicates, and 10,033 studies were uploaded to Covidence for screening. For articles with errata, addenda, and corrigenda, only one version was added to Covidence.

All titles and abstracts will first be screened in independent duplicate by two authors, with the other author resolving any conflicts. Abstracts which do not clearly meet the inclusion or exclusion criteria will be progressed to full text screening. Full text screening will be conducted in independent duplicate by two authors, with the other author resolving any conflicts for inclusion.

### Data charting

Data charting will also be conducted on Covidence. After full text screening, 5% of eligible articles will be charted in independent duplicate to evaluate charting consistency and to refine the Covidence data extraction template. The remaining articles will then be divided for individual extraction by the three authors. Advice from experts on anti-racism and neuroethics were sought to determine critical information to be extracted. Given that this scoping review primarily aims to audit reporting of RACE and diversity of research participants, the focus of extraction will not be on detailed study findings. Charting will be performed in two rounds. The first round will chart all eligible publications and will collect the following information:

1.) Reporting of RACE-ial (race; ancestry; country of birth; country of residence and migration/refugee status; culture, including language and religion; and/or ethnicity) information of the research participants
2.) Condition evaluated in the research
2.) Study objectives as indicated in the abstract
3.) Study methods as indicated in the abstract
4.) E-mail address of the corresponding author
5.) Journal
6.) Year of publication The second round will only involve articles that reported RACE-ial information. In addition to data collected during the first round, it will also collect:
7.) The RACE-ial variables collected and the terms used
8.) The categories used for the RACE-ial variables (e.g., Oceanian, North-west European, Southern and Eastern European, Northeast Asian, Southern and Central Asian, Southeast Asian, People of the Americas, North African and Middle Eastern, and Sub-Saharan African for ethnicity)
9.) Proportion of participants from different RACE-ial groups
10.) How racial/ethnic data was used in the publication (reported in a demographic table, used as a covariate, used to compare outcomes across different groups, etc.)
11.) Study objectives as indicated in the Introduction
12.) Study findings as indicated in the abstract
13.) Type and extent of consumer/patient engagement or leadership
14.) Discussion of potential impacts of RACE-ial variables on health outcomes, status, or access
15.) Recruitment efforts for particular RACE-ial minority groups
15.) Names and/or e-mail addresses of the lead and last authors
16.) Affiliations and location of the corresponding author
17.) Study context and/or participant location
18.) Bio/databank used (if applicable)
18.) Reporting of other demographic information (sex/gender, socio-economic status, etc.).

### Data synthesis

Research findings will be presented descriptively, with key findings on RACE reporting presented in tables and/or graphs and summarised using a narrative approach. Descriptive statistics will be used to compare reporting of RACE across the four conditions, different research methods/designs (based on study objectives), study locations, and period/year of publication.

## ETHICS AND DISSEMINATION

This review does not involve active human participation or unpublished secondary data, therefore approval from a human research ethics committee is not required.

Scoping review findings will be disseminated through conference presentations, social/professional media, university webpages, professional networks, and publication in a medical/health journal.

### Patient and public involvement

This publication analyses existing research studies, and therefore, involves no patients or members of the public. However, workshops with researchers and health practitioners, neuroethicists, and people with lived experience will be conducted to disseminate the review’s findings and collaboratively reflect on its implications for research and healthcare.

### IMPLICATIONS

Findings from this scoping review will provide a contemporary picture of demographic reporting and RACE-ial diversity in Australian BMH research. Focusing on four conditions (PTSD, depression, stroke, and dementia) would facilitate comparison of opportunities and challenges in diversifying research participants in neurologic, psychiatric, and neurovascular research. Moreover, including original research publications, regardless of the empirical method employed, would enable a nuanced understanding of barriers that are faced by different types of research. Findings on diversity in research across different Australian states and territories could highlight specific challenges faced by particular geographical contexts, given their demographics, history, and contemporary inter-racial/ethnic relations.

Insights and data from this review will inform subsequent phases of the larger project on anti-racist neuroethics for epistemic justice in BMH research (DE240100386). The list of lead and corresponding authors will be used to invite researchers for surveys, interviews, and workshops. The overall findings will be used to develop pragmatic recommendations for equitably increasing participant diversity in BMH research. With the increasing burden of BMH conditions; continuing disparities and racism experienced by particular populations; and growing anti-migrant, anti-refugee, and anti-Indigenous movements worldwide, it is imperative to ensure that health research accounts for the bodies, needs, and voices of diverse communities. Only through inclusive and equitable research practices can we truly safeguard mental health and well-being for all. No one should get left out and left behind.

## Data Availability

All data produced in the present study are available upon reasonable request to the authors.

## Acknowledgements

We acknowledge this year’s (2025) inputs, advice, and support from the following colleagues and collaborators for the scoping review and the overall project (DE240100386): Dr. Sarah Bourke (The Australian National University), Prof. Henry Brodaty (University of New South Wales), Prof. Julia Brown (University of California San Francisco), Melinda Burrows (The Australian National University), Prof. Laura Cabrera (Pennsylvania State University), Prof. Adrian Carter (Monash University), Prof. Jane Desborough (The Australian National University), Prof. Kevin Dunn (Western Sydney University), Prof. Kathomi Gatwiri (Southern Cross University), Prof. Frederic Gilbert (University of Tasmania), Prof. Kate Henne (The Australian National University), Dr. Samiul Hossain (Murdoch University), Prof Judy Illes (University of British Columbia), Prof. Lee-Fay Low (University of Sydney), Prof. Nicole Martinez-Martin (Stanford University), Dr. Christopher Mayes (Deakin University), Prof. Merryn McKinnon (The Australian National University), Dr. Bernard Saliba (University of Technology Sydney), and Prof. James Vickers (University of Tasmania)

## References

1. Canevelli M, Bruno G, Grande G, et al. Race reporting and disparities in clinical trials on Alzheimer’s disease: a systematic review. Neurosci Biobehav Rev 2019;101:122–28. doi: 10.1016/j.neubiorev.2019.03.020

2. Akinhanmi MO, Biernacka JM, Strakowski SM, et al. Racial disparities in bipolar disorder treatment and research: a call to action. Bipolar Disord 2018;20(6):506–14. doi: 10.1111/bdi.12638

3. Peterson RE, Kuchenbaecker K, Walters RK, et al. Genome-wide association studies in ancestrally diverse populations: opportunities, methods, pitfalls, and recommendations. Cell 2019;179(3):589–603. doi: 10.1016/j.cell.2019.08.051

4. Webb EK, Etter JA, Kwasa JA. Addressing racial and phenotypic bias in human neuroscience methods. Nat Neurosci 2022;25(4):410–14. doi: 10.1038/s41593-022-01046-0

5. Roberts SO, Bareket-Shavit C, Dollins FA, et al. Racial inequality in psychological research: trends of the past and recommendations for the future. Perspect Psychol Sci 2020;15(6):1295–309. doi: 10.1177/1745691620927709

6. Del Pozo B, Rich JD. Addressing racism in medicine requires tackling the broader problem of epistemic injustice. Am J Bioeth 2021;21(2):90–93. doi: 10.1080/15265161.2020.1861367

7. Australian Bureau of Statistics. Snapshot of Australia: a picture of the economic, social and cultural make-up of Australia on census night, 10 August 2021. Canberra (AU): ABS; 2022 [updated 2022 Oct 12; cited 2025 Apr 24]. Available from: https://www.abs.gov.au/statistics/people/people-and-communities/snapshot-australia/2021

8. Lewis C, Cohen PR, Bahl D, et al. Race and ethnic categories: a brief review of global terms and nomenclature. Cureus 2023;15(7):e41253. doi: 10.7759/cureus.41253

9. Suyemoto KL, Curley M, Mukkamala S. What do we mean by “ethnicity” and “race”? A consensual qualitative research investigation of colloquial understandings. Genealogy 2020; 4(3).

10. Pham TTL, Berecki-Gisolf J, Clapperton A, et al. Definitions of Culturally and Linguistically Diverse (CALD): A literature review of epidemiological research in Australia. Int J Environ Res Public Health 2021;18(2) doi: 10.3390/ijerph18020737

11. Australian Bureau of Statistics. National study of mental health and wellbeing: summary statistics on key mental health issues including national and state and territory estimates of prevalence of mental disorders. Canberra (AU): ABS; 2023.

12. Australian Institute of Health and Welfare. Australian burden of disease study 2024. Canberra (AU): AIHW; 2024.

13. Australian Institute of Health and Welfare. Heart, stroke and vascular disease: Australian facts. Canberra (AU): AIHW; 2024.

14. Australian Institute of Health and Welfare. Dementia in Australia. Canberra (AU): AIHW; 2024.

15. Schofield D, Cunich M, Shrestha R, et al. Indirect costs of depression and other mental and behavioural disorders for Australia from 2015 to 2030. BJPsych Open 2019;5(3):e40. doi: 10.1192/bjo.2019.26

16. Brown L, Hansnata E, La HA. Economic cost of dementia in Australia: 2016-2056. Canberra (AU): Alzheimer’s Australia, 2017.

17. Roberts AL, Gilman SE, Breslau J, et al. Race/ethnic differences in exposure to traumatic events, development of post-traumatic stress disorder, and treatment-seeking for post-traumatic stress disorder in the United States. Psychol Med 2011;41(1):71–83. doi: 10.1017/S0033291710000401

18. De Jesus-Romero R, Holder-Dixon AR, Buss JF, et al. Race, ethnicity, and other cultural background factors in trials of internet-based cognitive behavioral therapy for depression: Systematic Review. J Med Internet Res 2024;26:e50780. doi: 10.2196/50780

19. Lazaar N, Van Beek SE, Rirash AF, et al. Diversity in United States dementia prevention trials: an updated systematic review of eligibility criteria and recruitment strategies. Dement Geriatr Cogn Disord 2025 doi: 10.1159/000543905

20. Nanavati HD, Andrabi M, Arevalo YA, et al. Disparities in race and ethnicity reporting and representation for clinical trials in stroke: 2010 to 2020. J Am Heart Assoc 2024;13(6):e033467. doi: 10.1161/JAHA.123.033467

21. Towfighi A, Boden-Albala B, Cruz-Flores S, et al. Strategies to reduce racial and ethnic inequities in stroke preparedness, care, recovery, and risk factor control: a scientific statement from the American Heart Association. Stroke 2023;54(7):e371–e88. doi: 10.1161/STR.0000000000000437

22. Low LF, Barcenilla-Wong AL, Brijnath B. Including ethnic and cultural diversity in dementia research. Med J Aust 2019;211(8):345–46 e1. doi: 10.5694/mja2.50353

23. Minas H, Klimidis S, Kokanovic R. Depression in multicultural Australia: policies, research and services. Australia and New Zealand health policy 2007; 4.

24. Lynch E, Laver K, Levy T, et al. ‘The way that we are collecting and using data has evolved’ evaluating the Australian National Stroke Audit programme to inform strategic direction. BMJ Open Qual 2023;12(1) doi: 10.1136/bmjoq-2022-002136

25. Australian Institute of Health and Welfare. Size and sources of the health gap for Australia’s First Nations people 2017–2019. Canberra (AU): AIHW; 2024.

26. Davey RX. Health Disparities among Australia’s remote-dwelling Aboriginal people: a report from 2020. J Appl Lab Med 2021;6(1):125–41. doi: 10.1093/jalm/jfaa182

27. O’Brien P, Bunzli S, Lin I, et al. Addressing surgical inequity for Aboriginal and Torres Strait Islander people in Australia’s universal health care system: a call to action. ANZ J Surg 2021;91(3):238–44. doi: 10.1111/ans.16557

28. Australian Institute of Health and Welfare. Health of refugees and humanitarian entrants in Australia. Canberra (AU): AIHW; 2023.

29. Scanlon B, Brough M, Wyld D, et al. Equity across the cancer care continuum for culturally and linguistically diverse migrants living in Australia: a scoping review. Global Health 2021;17(1):87. doi: 10.1186/s12992-021-00737-w

30. Gatwiri K, Rotumah D, Rix E. BlackLivesMatter in healthcare: racism and implications for health inequity among Aboriginal and Torres Strait Islander peoples in Australia. Int J Environ Res Public Health 2021;18(9) doi: 10.3390/ijerph18094399

31. Gover AR, Harper SB, Langton L. Anti-Asian hate crime during the COVID-19 pandemic: exploring the reproduction of inequality. Am J Crim Justice 2020;45(4):647–67. doi: 10.1007/s12103-020-09545-1

32. Kamp A, Rachel S, Matteo V, et al. Asian Australian’s experiences and reporting of racism during the COVID-19 pandemic. J Intercult Stud 2024;45(3):452–72. doi: 10.1080/07256868.2023.2290676

33. Chibanda D, Jack HE, Langhaug L, et al. Towards racial equity in global mental health research. Lancet Psychiatry 2021;8(7):553–55. doi: 10.1016/S2215-0366(21)00153-X

34. Jimenez DE, Park M, Rosen D, et al. Centering culture in mental health: differences in diagnosis, treatment, and access to care among older people of color. Am J Geriatr Psychiatry 2022;30(11):1234–51. doi: 10.1016/j.jagp.2022.07.001

35. Niles PM, Jun J, Lor M, et al. Honoring Asian diversity by collecting Asian subpopulation data in health research. Res Nurs Health 2022;45(3):265–69. doi: 10.1002/nur.22229

36. Mervis J. Trump orders cause chaos at science agencies. Science 2025 2025 Feb 5.

37. Salles A, Banerjee AT, Caceres W, et al. Why and how academic medicine must champion diversity, equity, inclusion, and accessibility. Lancet 2025 doi: 10.1016/S0140-6736(25)00575-6

38. Cassidy C. Australian university researchers told ‘woke gender ideology’ among reasons behind Trump funding cuts. The Guardian 2025 2025 Mar 20.

39. Elias A, Paradies Y. Estimating the mental health costs of racial discrimination. BMC Public Health 2016;16(1):1205. doi: 10.1186/s12889-016-3868-1

40. Paradies Y, Ben J, Denson N, et al. Racism as a determinant of health: a systematic review and meta-analysis. PLOS ONE 2015;10(9):e0138511. doi: 10.1371/journal.pone.0138511

41. Fricker M. Epistemic injustice: power and the ethics of knowing: Oxford University Press, 2007.

42. Bhakuni H, Abimbola S. Epistemic injustice in academic global health. Lancet Glob Health 2021;9(10):e1465–e70. doi: 10.1016/S2214-109X(21)00301-6

43. Irzik G, Kurtulmus F. Distributive epistemic justice in science. Br J Philos Sci 2024;75(2):325–45.

44. Kowal E, Franklin H, Paradies Y. Reflexive antiracism: A novel approach to diversity training. Ethnicities 2013;13(3):316–37. doi: 10.1177/1468796812472885

45. Nelson J, Dunn K. Neoliberal anti-racism: Responding to ‘everywhere but different’ racism. Prog Hum Geogr 2016;41(1):26–43. doi: 10.1177/0309132515627019

46. Scholz B. We have to set the bar higher: towards consumer leadership, beyond engagement or involvement. Aust Health Rev 2022;46(4):509–12. doi: 10.1071/AH22022

47. Desborough J, Parkinson A, Lewis F, et al. A framework for involving coproduction partners in research about young people with type 1 diabetes. Health Expect 2022;25(1):430–42. doi: 10.1111/hex.13403

48. Tricco AC, Lillie E, Zarin W, et al. PRISMA extension for scoping reviews (PRISMA-ScR): checklist and explanation. Ann Intern Med 2018;169(7):467–73. doi: 10.7326/M18-0850

49. Peters MD, Godfrey CM, Khalil H, et al. Guidance for conducting systematic scoping reviews. Int J Evid Based Healthc 2015;13(3):141–6. doi: 10.1097/XEB.0000000000000050

50. Cardona LG, Yang MCL, Seon Q, et al. The methods of improving cultural sensitivity of depression scales for use among global indigenous populations: a systematic scoping review. Camb Prism Glob Ment Health 2023;10 doi: 10.1017/gmh.2023.75

51. Ostinelli EG, Zangani C, Giordano B, et al. Depressive symptoms and depression in individuals with internet gaming disorder: A systematic review and meta-analysis. J Affect Disord 2021;284:136–42. doi: 10.1016/j.jad.2021.02.014

52. Cheyne JD. Search strategy for retrieval of references on stroke healthcare in MEDLINE Ovid. In: Cochrane Stroke Group CoMaVM, University of Edinburgh,, ed., 2020.

53. Wardlaw JM, Murray V, Berge E, et al. Recombinant tissue plasminogen activator for acute ischaemic stroke: an updated systematic review and meta-analysis. Lancet 2012;379(9834):2364–72. doi: 10.1016/S0140-6736(12)60738-7

54. Coventry PA, Meader N, Melton H, et al. Psychological and pharmacological interventions for posttraumatic stress disorder and comorbid mental health problems following complex traumatic events: Systematic review and component network meta-analysis. PLoS Med 2020;17(8):e1003262. doi: 10.1371/journal.pmed.1003262

55. Cheyne JD. Search strategies for retrieval of references for effective interventions to support the resilience and mental health of frontline health and social care staff during a global health crisis and following de-escalation. In: Cochrane Stroke Group CoMaVM, University of Edinburgh,, ed., 2020.

56. Bellaver B, Ferrari-Souza JP, Uglione da Ros L, et al. Astrocyte biomarkers in alzheimer disease: a systematic review and meta-analysis. Neurology. 2021;96(24):e2944–e55. doi: 10.1212/WNL.0000000000012109

57. Wallace L, Theou O, Rockwood K, et al. Relationship between frailty and Alzheimer’s disease biomarkers: a scoping review. Alzheimers Dement (Amst) 2018;10:394–401. doi: 10.1016/j.dadm.2018.05.002

58. Maharaj R, Ndwiga D, Chutiyami M. Mental health and wellbeing of international students in Australia: a systematic review. J Ment Heal 2024 doi: 10.1080/09638237.2024.2390393

59. Lirios A, Mullens AB, Daken K, et al. Sexual and reproductive health literacy of culturally and linguistically diverse young people in Australia: a systematic review. Cult Health Sex 2024;26(6):790–807. doi: 10.1080/13691058.2023.2256376

60. Grant MJ, Booth A. A typology of reviews: an analysis of 14 review types and associated methodologies. Health Information & Libraries Journal 2009;26(2):91–108. doi: 10.1111/j.1471-1842.2009.00848.x

61. Ong DS, von Mollendorf C, Mulholland K, et al. Measles seroprevalence in infants under nine months of age in low- and middle-income countries: a systematic review and meta-analysis. J Infect Dis 2025 doi: 10.1093/infdis/jiaf177

62. Bramer WM, Rethlefsen ML, Kleijnen J, et al. Optimal database combinations for literature searches in systematic reviews: a prospective exploratory study. Syst Rev 2017;6(1):245. doi: 10.1186/s13643-017-0644-y

63. Ryan JC, Viana JN, Sellak H, et al. Defining precision health: a scoping review protocol. BMJ Open 2021;11(2):e044663. doi: 10.1136/bmjopen-2020-044663

64. Viana JN, Edney S, Gondalia S, et al. Trends and gaps in precision health research: a scoping review. BMJ Open 2021;11(10):e056938. doi: 10.1136/bmjopen-2021-056938

