## Appendix 1 for "Reporting of RACE in Australian brain and mental health research: protocol for a scoping review and diversity audit"

### Embase Search Strings

| Description | | Search string |
|---|---|---|
| Post-traumatic stress disorder (PTSD) related terms | 1. | (exp posttraumatic stress disorder/) or ((PTSD).ti,ab,kw.) or ((posttrauma$ or post-trauma$ or post traumatic stress).ti,ab,kw.) or (((trauma$) adj3 (disorder or neurosis or psychos$ or syndrome or expos$ or event$ or experienc$)).ti,ab,kw.) or ((traumatic stress or traumatic memor$ or traumati?ation).ti,ab,kw.)).ti,ab,kw.) |
| Depression-related terms | 2. | (exp depressive disorder/) or (exp depression/) or (exp dysthymic disorder/) or ((depress* or dysthymi*).ti,ab,kw.) |
| Stroke-related terms | 3. | (exp cerebrovascular disease/ or exp basal ganglia cerebrovascular disease/ or exp brain ischemia/ or exp brain infarction/ or exp transient ischemic attack/ or exp vertebrobasilar insufficiency/ or exp carotid artery disease/ or carotid artery thrombosis/ or exp cerebral small vessel disease/ or exp cerebral amyloid angiopathy/ or exp lacunar stroke/ or exp cerebrovascular trauma/ or exp vertebral artery dissection/ or exp intracranial artery disease/ or exp cerebral artery disease/ or exp cerebral amyloid angiopathy/ or exp anterior cerebral artery infarction/ or exp middle cerebral artery infarction/ or exp posterior cerebral artery infarction/ or exp moyamoya disease/ or exp intracranial aneurysm/ or exp intracranial arteriosclerosis/ or exp intracranial arteriovenous malformation/ or exp intracranial embolism/ or exp intracranial thrombosis/ or exp intracranial hemorrhage/ or exp cerebral hemorrhage/ or exp hypertensive intracranial hemorrhage/ or exp subarachnoid hemorrhage/ or exp stroke/ or exp intracranial vasospasm/ or exp brain hypoxia-ischemia/) or ((stroke$ or apoplex$ or cerebral vasc$ or cerebrovasc$ or cva or transient isch?emic attack$ or tia$).ti,ab,kw.) or (((brain or cerebr$ or cerebell$ or vertebrobasil$ or hemispher$ or intracran$ or intracerebral or infratentorial or supratentorial or middle cerebr$ or mca$ or anterior circulation).ti,ab,kw.) and ((isch?emi$ or infarct$ or thrombo$ or emboli$ or occlus$ or hypoxi$).ti,ab,kw.)) |
| Dementia-related terms | 4. | (exp dementia/) or (dementia* or "frontotemporal dementia" or "frontotemporal lobar degeneration" or "behavioral variant frontotemporal dementia" or Lewy* or DLB or LBD or ATD or FAD or EOAD or “Alzheimer’s” or PDD or “Parkinson’s disease dementia” or “Parkinson’s dementia” or "vascular dementia" or "arteriosclerotic dementia" or “dementia multi-infarct”).ti,ab,kw. |
| Combination of disease-related terms | 5. | 1 or 2 or 3 or 4 |
| Terms for studies in Australia | 6. | exp Australia/ |
| | 7. | (Adelaide or Brisbane or Canberra or Darwin or Hobart or Melbourne or Perth or Sydney or “South Australia” or Queensland or “Australian Capital Territory” or “Northern Territory” or Tasmania or Victoria or “Western Australia” or “New South Wales”).ti,ab,kw. |
| | 8. | 6 or 7 |

| | | |
|---|---|---|
| Research on the four conditions in Australia | 9. | 5 and 8 |
| Research published on 1 January 2020 to 31 December 2024 | 10. | limit 9 to yr="2020-2024" |
| Terms to exclude reviews and meta-analyses | 11. | (“systematic review$” OR “scoping review$” OR “narrative review$” OR “rapid review$” OR “literature review$” OR “integrative review$” OR “methodological review$” OR “comparative effectiveness review$” OR “prognostic review$” OR “psychometric review$” OR “review of economic evaluation$” OR “umbrella review$” OR “meta analy*” OR “meta-analy*” OR “critical review$” OR “mapping review$” OR “mixed stud* review$” OR “mixed methods review$” OR “evidence synthesis” OR “state-of-the-art review$” OR “systematic search and review$” OR “systemati$ed review$”).ti,ab,kw. |
| Terms to exclude animal studies | 12. | (exp animals/ OR (rat$ OR mouse OR mice OR rodent$ OR swine$ OR porcine$ OR murine$ OR sheep$ OR lamb$ OR pig$ OR piglet$ OR rabbit$ OR cat$ OR dog$ OR cattle OR bovine OR monkey$ OR trout OR marmoset$ OR macaque$ OR zebrafish OR “fruit fl*” OR roundworm).ti,ab,kw.) not human*.sh. |
| Final search string | 25. | 10 not (11 or 12) |

### CINAHL Search Strings

| Description | | Search string |
|---|---|---|
| Search mode: Proximity | | |
| Post-traumatic stress disorder (PTSD) related terms | 1. | “posttraumatic stress disorder” or PTSD or posttrauma$ or post-trauma$ or “post traumatic stress” or (trauma$ N3 (disorder or neurosis or psychos$ or syndrome)) or “traumatic stress” or “traumatic memor$” or traumati?ation or (trauma$ N3 (expos$ or event$ or experienc$)) |
| Depression-related terms | 2. | “depressive disorder” or depression or “dysthymic disorder” or depress* or dysthymi* |
| Stroke-related terms | 3. | (“cerebrovascular disease” or “basal ganglia cerebrovascular disease” or “brain ischemia” or “brain infarction” or “transient ischemic attack” or “vertebrobasilar insufficiency” or “carotid artery disease” or “carotid artery thrombosis” or “cerebral small vessel disease” or “cerebral amyloid angiopathy” or “lacunar stroke” or “cerebrovascular trauma” or “vertebral artery dissection” or “intracranial artery disease” or “cerebral artery disease” or “cerebral amyloid angiopathy” or “anterior cerebral artery infarction” or “middle cerebral artery infarction” or “posterior cerebral artery infarction” or “moyamoya disease” or “intracranial aneurysm” or “intracranial arteriosclerosis” or “intracranial arteriovenous malformation” or “intracranial embolism” or “intracranial thrombosis” or “intracranial hemorrhage” or “cerebral hemorrhage” or “hypertensive intracranial hemorrhage” or “subarachnoid hemorrhage” or “stroke” or “intracranial vasospasm” or “brain hypoxia-ischemia” or “stroke$” or “apoplex$” or “cerebral vasc$” or “cerebrovasc$” or “cva” or “transient isch?emic attack$” or “tia$”) or ((“brain” or “cerebr$” or “cerebell$” or “vertebrobasil$” or “hemispher$” or “intracran$” or “intracerebral” or “infratentorial” or “supratentorial” or “middle cerebr$” or “mca$” or “anterior circulation”) and (“isch?emi$” or “infarct$” or “thrombo$” or “emboli$” or “occlus$” or “hypoxi$”)) |
| Dementia-related terms | 4. | dementia or dementia* or "frontotemporal dementia" or "frontotemporal lobar degeneration" or "behavioral variant frontotemporal dementia" or Lewy* or DLB or LBD or ATD or FAD or EOAD or “Alzheimer’s” or PDD or “Parkinson’s disease dementia” or “Parkinson’s dementia” or "vascular dementia" or "arteriosclerotic dementia" or “dementia multi-infarct” |
| Combination of disease-related terms | 5. | S1 or S2 or S3 or S4 |
| Terms for studies in Australia | 6. | Australia |
| | 7. | Adelaide or Brisbane or Canberra or Darwin or Hobart or Melbourne or Perth or Sydney or “South Australia” or Queensland or “Australian Capital Territory” or “Northern Territory” or Tasmania or Victoria or “Western Australia” or “New South Wales” |
| | 8. | S6 or S7 |
| Research on the four conditions in Australia | 9. | S5 and S8 |
| Terms to exclude reviews and meta-analyses | 10. | “systematic review$” OR “scoping review$” OR “narrative review$” OR “rapid review$” OR “literature review$” OR “integrative review$” OR “methodological review$” OR “comparative effectiveness review$” OR |

| | | |
|---|---|---|
| | | “prognostic review$” OR “psychometric review$” OR “review of economic evaluation$” OR “umbrella review$” OR “meta analy*” OR “meta-analy*” OR “critical review$” OR “mapping review$” OR “mixed stud* review$” OR “mixed methods review$” OR “evidence synthesis” OR “state-of-the-art review$” OR “systematic search and review$” OR “systemati$ed review$” |
| Terms to exclude animal studies | 11. | (animal$ OR rat$ OR mouse OR mice OR rodent$ OR swine$ OR porcine$ OR murine$ OR sheep$ OR lamb$ OR pig$ OR piglet$ OR rabbit$ OR cat$ OR dog$ OR cattle OR bovine OR monkey$ OR trout OR marmoset$ OR macaque$ OR zebrafish OR “fruit fl*” OR roundworm) NOT human$ |
| Final search string | 12. | S9 not (S10 or S11)<br><br>limit from 2020-2024 |

### Scopus Search Strings

| Description | Search string |
|---|---|
| Post-traumatic stress disorder (PTSD) related terms | ( (TITLE-ABS-KEY ( "stress disorders" OR "post traumatic" OR ptsd OR posttrauma* OR "traumatic stress" OR "traumatic memor$" OR traumati?ation OR ( trauma* W/2 ( disorder OR neurosis OR psycho* OR syndrome OR expos* OR event* OR experienc* ) ) )<br><br>OR |
| Depression-related terms | TITLE-ABS-KEY ( depress* OR dysthymi* OR melancholi* )<br><br>OR |
| Stroke-related terms | TITLE-ABS-KEY ( "cerebrovascular disease" OR "brain ischemia" OR "brain infarction" OR "transient ischemic attack" OR "vertebrobasilar insufficiency" OR "carotid artery disease" OR "carotid artery thrombosis" OR "cerebral small vessel disease" OR "cerebral amyloid angiopathy" OR "lacunar stroke" OR "cerebrovascular trauma" OR "vertebral artery dissection" OR "intracranial artery disease" OR "cerebral artery disease" OR "cerebral amyloid angiopathy" OR "anterior cerebral artery infarction" OR "middle cerebral artery infarction" OR "posterior cerebral artery infarction" OR "moyamoya disease" OR ( intracranial PRE/0 ( aneurysm OR arteriosclerosis OR "arteriovenous malformation" OR embolism OR thrombosis OR hemorrhage ) ) OR ( ( cerebral OR "hypertensive intracranial" OR subarachnoid ) PRE/0 hemorrhage ) OR "stroke" OR "intracranial vasospasm" OR "brain hypoxia-ischemia" OR "stroke$" OR "apoplex$" OR "cerebral vasc$" OR "cerebrovasc$" OR "cva" OR "transient isch?emic attack$" OR "tia$" OR ( ( "brain" OR "cerebr$" OR "cerebell$" OR "vertebrobasil$" OR "hemispher$" OR "intracran$" OR "intracerebral" OR "infratentorial" OR "supratentorial" OR "middle cerebr$" OR "mca$" OR "anterior circulation" ) AND ( "isch?emi$" OR "infarct$" OR "thrombo$" OR "emboli$" OR "occlus$" OR "hypoxi$" ) ) )<br><br>OR |
| Dementia-related terms | TITLE-ABS-KEY ( dementia* OR "Frontotemporal lobar degeneration" OR lewy* OR dlb OR lbd OR atd OR fad OR eoad OR "Alzheimer's" OR pdd ) )<br><br>AND |
| Terms for studies in Australia | TITLE-ABS-KEY ( Australia OR “Adelaide” OR “Brisbane” OR “Canberra” OR “Darwin” OR “Hobart” OR “Melbourne” OR “Perth” OR “Sydney” OR “South Australia” OR “Queensland” OR “Australian Capital Territory” OR “Northern Territory” OR “Tasmania” OR “Victoria” OR “Western Australia” OR “New South Wales” )<br><br>AND NOT |
| Terms to exclude reviews and meta-analyses | TITLE-ABS-KEY ( "systematic review$" OR "scoping review$" OR "narrative review$" OR "rapid review$" OR "literature review$" OR "integrative review$" OR "methodological review$" OR "comparative effectiveness review$" OR "prognostic review$" OR "psychometric review$" OR "review of economic evaluation$" OR "umbrella review$" OR "meta analy*" OR "meta-analy*" OR "critical review$" OR "mapping review$" OR "mixed stud* review$" OR "mixed methods review$" OR "evidence synthesis" OR "state-of-the-art review$" OR "systematic search and review$" OR "systemati$ed review$" ) |

| | |
|---|---|
| | AND NOT |
| Terms to exclude animal studies | TITLE-ABS-KEY ( ( animal$ OR rat$ OR mouse OR mice OR rodent$ OR swine$ OR porcine$ OR murine$ OR sheep$ OR lamb$ OR pig$ OR piglet$ OR rabbit$ OR cat$ OR dog$ OR cattle OR bovine OR monkey$ OR trout OR marmoset$ OR macaque$ OR zebrafish OR "fruit fl*" OR roundworm ) not AND human* ) ) |
| | AND |
| Research published on 1 January 2020 to 31 December 2024 | PUBYEAR > 2019 AND PUBYEAR < 2025 |

### Web of Science

| Description | | Search string |
|---|---|---|
| Post-traumatic stress disorder (PTSD) related terms | 1. | TS=((“posttraumatic stress disorder” OR “PTSD” OR “posttrauma$” OR “post-trauma$” OR “post traumatic stress” OR “traumatic disorder” OR “traumatic neurosis” OR “traumatic psychos$” OR “traumatic syndrome” OR “traumatic stress” OR “traumatic memor$” OR “traumati?ation” OR (trauma$ NEAR/3 (disorder OR neurosis OR psychos$ OR syndrome OR expos$ OR event$ OR experience$)) OR |
| Depression-related terms | | (“depressive disorder” OR “depression” OR “dysthymic disorder” OR “depress$” OR “dysthmi$”) OR |
| Stroke-related terms | | ((“cerebrovascular disease” OR “basal ganglia cerebrovascular disease” OR “brain ischemia” OR “brain infarction” OR “transient ischemic attack” OR “vertebrobasilar insufficiency” OR “carotid artery disease” OR “carotid artery thrombosis” OR “cerebral small vessel disease” OR “cerebral amyloid angiopathy” OR “lacunar stroke” OR “cerebrovascular trauma” OR “vertebral artery dissection” OR “intracranial artery disease” OR “cerebral artery disease” OR “cerebral amyloid angiopathy” OR “anterior cerebral artery infarction” OR “middle cerebral artery infarction” OR “posterior cerebral artery infarction” OR “moyamoya disease” OR “intracranial aneurysm” OR “intracranial arteriosclerosis” OR “intracranial arteriovenous malformation” OR “intracranial embolism” OR “intracranial thrombosis” OR “intracranial hemorrhage” OR “cerebral hemorrhage” OR “hypertensive intracranial hemorrhage” OR “subarachnoid hemorrhage” OR “stroke” OR “intracranial vasospasm” OR “brain hypoxia-ischemia” OR “stroke$” OR “apoplex$” OR “cerebral vasc$” OR “cerebrovasc$” OR “cva” OR “transient isch?emic attack$” OR “tia$”) OR ((“brain” OR “cerebr$” OR “cerebell$” OR “vertebrobasil$” OR “hemispher$” OR “intracran$” OR “intracerebral” OR “infratentorial” OR “supratentorial” OR “middle cerebr$” OR “mca$” OR “anterior circulation”) AND (“isch?emi$” OR “infarct$” OR “thrombo$” OR “emboli$” OR “occlus$” OR “hypoxi$”))) OR |
| Dementia-related terms | | (“dementia” OR “dementia*” OR “frontotemporal dementia” OR “frontotemporal lobar degeneration” OR “behavioral variant frontotemporal dementia” OR “Lewy*” OR “DLB” OR “LBD” OR “ATD” OR “FAD” OR “EOAD” OR “Alzheimer’s” OR “PDD” OR “Parkinson’s disease dementia” OR “Parkinson’s dementia” OR “vascular dementia” OR “arteriosclerotic dementia” OR “dementia multi-infarct”) ) ) |
| Terms for studies in Australia | 2. | TS=(“Australia” OR “Adelaide” OR “Brisbane” OR “Canberra” OR “Darwin” OR “Hobart” OR “Melbourne” OR “Perth” OR “Sydney” OR “South Australia” OR “Queensland” OR “Australian Capital Territory” OR “Northern Territory” OR “Tasmania” OR “Victoria” OR “Western Australia” OR “New South Wales” ) |
| Research on the four conditions in Australia | 3. | #1 AND #2 |
| Terms to exclude reviews and meta-analyses | 4. | TS=(“systematic review$” OR “scoping review$” OR “narrative review$” OR “rapid review$” OR “literature review$” OR “integrative review$” OR “methodological review$” OR “comparative effectiveness review$” OR “prognostic review$” OR “psychometric review$” OR “review of economic evaluation$” OR “umbrella review$” OR “meta analy” OR “meta-analy” OR “critical review$” OR “mapping review$” OR “mixed |

| | | |
|---|---|---|
| | | stud* review$" OR "mixed methods review$" OR "evidence synthesis" OR "state-of-the-art review$" OR "systematic search and review$" OR "systematised review$" OR "systematized review$") |
| Terms to exclude animal studies | 5. | TS=("animals" OR "rat$" OR "mouse" OR "mice" OR "rodent$" OR "swine$" OR "porcine$" OR "murine$" OR "sheep$" OR "lamb$" OR "pig$" OR "piglet$" OR "rabbit$" OR "cat$" OR "dog$" OR "cattle" OR "bovine" OR "monkey$" OR "trout" OR "marmoset$" OR "macaque$" OR "zebrafish" OR "fruit fl*" OR "roundworm" ) |
| | 6. | NOT TS=(human$) |
| | 7. | (#4 AND #5) NOT #6 |
| Final search string | 8. | #3 NOT #7<br><br>limit from 2020-2024 |

### PsycInfo Search Strings

| Description | Search string |
|---|---|
| Post-traumatic stress disorder (PTSD) related terms | 1. exp Posttraumatic Stress Disorder /<br>2. (PTSD or posttrauma$ or post-trauma$ or “post traumatic stress” or “traumatic stress” or “traumatic memor$” or traumati?ation).ti,ab,id.<br>3. ((trauma$) adj3 (disorder or neurosis or psychos$ or syndrome or expos$ or event$ or experienc$)).ti,ab,id.<br>4. 1 or 2 or 3 |
| Depression-related terms | 5. exp depressive disorder/ OR exp "depression (emotion)"/ OR exp dysthymic disorder/<br>6. (depress* or dysthymi*).ti,ab,id.<br>7. 5 or 6 |
| Stroke-related terms | 8. exp ischemia/ OR exp Cerebrovascular Accidents/ OR exp Cerebrovascular Disorders/ OR exp Cerebral Hemorrhage/ OR exp Cerebral Small Vessel Disease/ or exp subarachnoid hemorrhage/<br>9. (stroke$ or apoplex$ OR cerebral vasc$ OR cerebrovasc$ OR cva OR “transient isch?emic attack$” OR “vertebrobasilar insufficiency” OR tia$ OR “carotid artery diseases” OR “carotid artery thrombosis” OR “cerebral amyloid angiopathy” OR “cerebrovascular trauma” OR “vertebral artery dissection” OR “intracranial arterial diseases” OR “cerebral arterial diseases” OR “cerebral amyloid angiopathy” OR “intracranial arteriovenous malformations” OR “intracranial aneurysm” OR “intracranial arteriosclerosis” OR “intracranial arteriovenous malformations” OR “moyamoya disease” OR “intracranial vasospasm” OR “hypoxia-ischemia”).ti,ab,id.<br>10. (brain or cerebr$ or cerebell$ or vertebrobasil$ or hemispher$ or intracran$ or intracerebral or “anterior cerebral artery” or “infratentorial or supratentorial” or “middle cerebr$” or mca$ or “anterior circulation” or “middle cerebral artery” or “posterior cerebral artery”).ti,ab,id.<br>11. (isch?emi$ or infarct$ or thrombo$ or emboli$ or occlus$ or hypoxi$ or hemorrh$).ti,ab,id.<br>12. 10 and 11<br>13. 8 or 9 or 12 |
| Dementia-related terms | 14. Exp Dementia/<br>15. (Dementia* or "Frontotemporal dementia" or "Frontotemporal lobar degeneration" or "behavioral variant frontotemporal dementia" or Lewy* or DLB or LBD or ATD or FAD or EOAD or “Alzheimer’s” or PDD or “Parkinson’s disease dementia” or “Parkinson’s dementia” or "vascular dementia" or "Arteriosclerotic Dementia" or “dementia multi-infarct”).ti,ab,id.<br>16. 14 or 15 |
| Combination of disease-related terms | 17. 4 or 7 or 13 or 16 |
| Terms for studies in Australia | 18. exp Australia/<br>19. (Adelaide or Brisbane or Canberra or Darwin or Hobart or Melbourne or Perth or Sydney or “South Australia” or Queensland or “Australian Capital Territory” or “Northern Territory” or Tasmania or Victoria or “Western Australia” or “New South Wales”).ti,ab,id.<br>20. 18 or 19 |

| | |
|---|---|
| Research on the four conditions in Australia | 21. 17 and 20 |
| Research published on 1 January 2020 to 31 December 2024 | 22. limit 21 to yr="2020-2024" |
| Terms to exclude reviews and meta-analyses | 23. ("systematic review$" OR "scoping review$" OR "narrative review$" OR "rapid review$" OR "literature review$" OR "integrative review$" OR "methodological review$" OR "comparative effectiveness review$" OR "prognostic review$" OR "psychometric review$" OR "review of economic evaluation$" OR "umbrella review$" OR "meta analy*" OR "meta-analy*" OR "critical review$" OR "mapping review$" OR "mixed stud* review$" OR "mixed methods review$" OR "evidence synthesis" OR "state-of-the-art review$" OR "systematic search and review$" OR "systemati$ed review$").ti,ab,id. |
| Terms to exclude animal studies | 24. (exp animals/ OR (rat$ OR mouse OR mice OR rodent$ OR swine$ OR porcine$ OR murine$ OR sheep$ OR lamb$ OR pig$ OR piglet$ OR rabbit$ OR cat$ OR dog$ OR cattle OR bovine OR monkey$ OR trout OR marmoset$ OR macaque$ OR zebrafish OR "fruit fl*" OR roundworm).ti,ab,id.) not human*.sh. |
| Final search string | 26. 22 not (23 or 24) |
